# Dynamic Training of a Novelty Classifier Algorithm for Real-Time Early Seizure Onset Detection

**DOI:** 10.1101/2021.03.06.21253045

**Authors:** Daniel Ehrens, Mackenzie C. Cervenka, Gregory K. Bergey, Christophe C. Jouny

## Abstract

The objective of this study was to develop an adaptive framework for seizure detection in real-time that is practical to use in the Epilepsy Monitoring Unit (EMU) as a warning signal, and whose output helps characterize epileptiform activity. Our framework uses a one-class Support Vector Machine (SVM) that is being trained dynamically according to past activity in all available channels. This is done to evaluate the novelty of the current instance according to previous activity. Our algorithm was tested on intracranial EEG from human epilepsy patients that are admitted to the EMU for presurgical evaluation. In this study, we compared multiple configurations for using a one-class SVM to assess if there is significance over specific neural features or electrode locations. Our results show our algorithm is capable of running in real-time and achieving a high performance for early seizure-onset detection with a low false-positive rate and robustness to different types of seizure-onset patterns as well as to the number of channels used. This algorithm offers a solution to warning systems in the EMU as well as a tool for seizure characterization during post-hoc analysis of intracranial EEG data for surgical resection of the epileptogenic network.

**Highlights:**

- This study proposes a dynamic training algorithm that efficiently detects sudden novel changes in intracranial electroencephalographic activity, creating a reliable seizure onset detection algorithm that does not need prior training.
- The algorithm described has the capability to be implemented in real-time, independently of the number of channels that are being analyzed.
- The presented detector shows high performance and reliability to be easily implemented in the Epilepsy Monitoring Unit to quickly alert clinical staff of seizure events.

## 1. Introduction

Epilepsy is a neurological condition that affects approximately 70 million people worldwide (Ngugi et al. 2011). About one-third of patients with epilepsy have drug refractory seizures and may be candidates for resective epilepsy surgery or deep brain stimulation therapy. Pre-surgical evaluation for any invasive alternative requires seizure localization through long-term video EEG-monitoring in epilepsy monitoring units (EMUs). This process requires coordination and can be resource intensive to ensure patient safety and analyze the high amount of EEG data collected. Failure to detect electrographic seizures can increase risk of physical and neuronal injury, Atkinson et al. reported in a 2008 study of 170 events in a single hospital, 44 clinical seizures and 57 electrographic seizures did not trigger a staff response (Atkinson et al. 2012). While many centers have a higher response rate, it remains likely that the implementation of automatized early seizure detection algorithms, would unequivocally improve patient safety and efficiency in the EMU (Noe and Drazkowski 2009; Jouny et al. 2011; Lee and Shah 2013; Kamitaki et al. 2019). Reliable early seizure-onset detection in real-time would provide a trustworthy warning system in the EMU to alert personnel and allow assessment of the patient during an ongoing seizure. Such algorithm must be highly sensitive and specific to seizure activity to avoid alarm fatigue (Bi et al. 2020), while having minimum detection latency (Jouny et al. 2011).

Although automatized seizure detection has been a research topic of renewed interest in light of development of innovative therapies including responsive neurostimulation, it still remains a challenge (Mormann et al. 2007; Freestone et al. 2015; Stacey 2018). The complexity of seizure dynamics, the variability of seizure-onset patterns and the localization of the seizure onset foci, can often limit the efficacy of seizure detection algorithms as well as their practicality for the EMU. The wide variety of patients with focal epilepsy admitted to the EMU complicates the task of designing an algorithm that is sensitive to each patient’s stereotypical ictal pattern(s) (Fürbass et al. 2015). Moreover, localization of the seizure onset foci is a key objective in pre-surgical evaluation. This often requires the implantation of a large number of intracranial electrodes to provide sufficient spatial information about the extent of the seizure onset zone. The large number of channels to analyze reduces the signal to noise ratio, making it more challenging for seizure detection algorithms to achieve high specificity which is vital for clinical use. An algorithm with a high false positive rate does not provide a reliable alarm, and is typically ignored by clinical staff (Lee and Shah 2013; Kamitaki et al. 2019; Bi et al. 2020).

Modern seizure detection algorithms make use of machine learning algorithms to classify electroencephalographic features into ictal or non-ictal events (Meier et al. 2008b; Kharbouch et al. 2011; Kuhlmann et al. 2018). The performance of some of these algorithms relies highly on their training data, which directly affects the performance of the classifier. The vast number of seizure onset patterns and variability in electrode locations makes it extremely challenging to build a training set that accurately represents the feature-space for all patients. Lastly, this is particularly difficult in the EMU when no prior ictal data is available for the specific patient.

Support Vector Machines (SVMs) are a general class of machine learning algorithms (Scholkopf, 1998) with several advantages over other machine learning techniques. The one-class SVM is a powerful algorithm for seizure detection proposed by Gardner et al. 2006. It was designed for unbalanced datasets, where normal samples are abundant, but abnormal samples are rare. This model is particularly well suited for seizure detection in which the ictal events are rare in comparison to the duration of the non-ictal state. Additionally the one-class SVM is practical to implement in real-time as it is computationally inexpensive compared to other novelty detection methods (Hoffmann 2007).This algorithm offers a framework to build an adaptive detection method that does not require prior ictal data, making it suitable for implementation in the EMU.

In this study, we used intracranial electrographical recordings from patients with focal epilepsy that were admitted for presurgical evaluation requiring intracranial recordings. We compare the use of one-class SVMs in three different configurations with the goal of assessing the overall performance of the algorithm in each configuration. Here, we introduce an innovative patient-specific seizure detection paradigm in which training of the classifier is performed ad-hoc, resulting in a dynamical and self-adapting algorithm. Our algorithm trains every second with a delayed moving window of interictal data making it patient-specific and free of prior training before being implemented in the EMU. Our results show the performance of the algorithm for real-time early seizure onset detection, as well as for universal generic seizure detection.

## 2. Methods

Our proposed algorithm performs a sliding window analysis of the EEG every second. Once the EEG is acquired, non-neural artifacts are removed. Features sensitive to seizure onset are extracted and preprocessed to train a one-class SVM and classify the novelty of the current instance. The output of the one-class SVM is then postprocessed to identify epileptiform activity and trigger the detection of an event in real-time. The algorithm has a half-second processing time from beginning to end, which is repeated every second. The results shown in this study are done post hoc, however our algorithm was designed to run in real-time (i.e. ad-hoc). All analyses were done using custom-written scripts in Matlab (The MathWorks Inc., Natick, MA, USA), on a Windows 64-bit machine with an Intel Core i7 3.6 GHz processor. LIBSVM was used for all SVM computations (Chang and Lin 2001).

### 2.1 Clinical data

Intracranial recordings from 35 consecutive patients admitted to the Johns Hopkins Hospital EMU for presurgical evaluation were used in this study. The patient cohort included 62% men and 38% women, with a mean age of 33.7±14.1 years. Patients were implanted with a combination of subdural grids, strips and/or depth electrodes to record ECoG and iEEG signals respectively; through the rest of the paper we will just refer to these as EEG. The dataset consists of a total of 655 hours of EEG recordings containing 132 seizures. Recorded seizures originated from different structures in the brain (temporal, frontal, parietal or occipital lobe). Seizure datasets included focal onset seizures with and without loss of awareness and focal to bilateral tonic-clonic activity. Seizure onset times were identified by certified epileptologists (M.C.C. and G.K.B.). For each patient we used an epoch of continuous EEG data ranging from 3 to 48 hours, with an average length of 19±11 hours. The epoch of continuous EEG contained in average 3.8 ictal events, ranging from 1 to 11 seizures. The number of channels recorded per patient ranged from 23 to 169, with an average of 80 channels. Channels that showed continuous artifact and were not used for clinical evaluation were excluded for this study. Included patients had at least two hours of continuous EEG recording before the first seizure. Patients were excluded if they only had electrographical seizures without clinical symptoms and/or did not have clear background activity. EEG data was digitized and stored using a Neurofax EEG-1200 Platform (Nihon Kohden, Tokyo, Japan) with a 2000 Hz sampling frequency. Our research protocol was reviewed and approved by the Johns Hopkins Institutional Review Board and data were stored in compliance with HIPAA regulations.

### 2.2 Artifact detection

EEG data was processed using a bipolar measurement between pairs of channels offering a better spatial localization than reference recordings (Nunez and Srinivasan 2006). The bipolar method also provides common-mode rejection, reducing line noise and artifacts common to all electrodes. Artifacts caused by non-neural sources were detected and removed from the raw EEG signal. We developed an artifact detection algorithm that extracts features sensitive to non-neural artifacts (Delorme et al. 2007). The features used for artifact detection were kurtosis, power in delta band [0 – 4 Hz], maximal voltage, and the first derivative of the signal. Features were extracted every second from the previous four seconds of EEG signal. The threshold for each feature was selected at 120% of the maximum value from a random set of 10 seizures. This was done to calibrate the algorithm and ensure ictal activity would not be identified as an artifact, which would affect the algorithm’s sensitivity. This was done as a one-time initialization of the algorithm. Once artifacts were detected, we store which channels had artifacts for artifact removal during the preprocessing stage.

### 2.3 Feature extraction

To characterize EEG properties, we used a set of 6 features that have already been characterized and shown to be useful for early seizure onset detection for temporal and extra temporal stereotypes of seizures (Jouny and Bergey 2012). Features were selected according to their sensitivity to seizure onset patterns and their computational practicality to be implemented in a real-time algorithm. The set of features included linear and spectral parameters, and complexity measures. The power in specific frequency bands (β - [12-30 Hz], γ - [30-80 Hz], high-frequencies - [80-200Hz]) were extracted using Welch’s method (Welch 1967). Complexity features included the Higuchi fractal dimension (Higuchi 1988) and the Lempel-Ziv complexity (Lempel and Ziv 1976). The linear feature, line-length was also extracted from the EEG, which has been described as an optimal feature for online low complexity detection (Logesparan et al., 2012).

### 2.4 Preprocessing

Once features were extracted from the EEG, preprocessing was completed for training of the SVM. This involved artifact filtering and normalization. Using the results from the artifact detector we replaced the values in the feature vector where artifacts were detected. The feature vector was patched using the average of the last minute. This replaced the contaminated feature vector with a value that would not be novel to the SVM. Additionally, if a specific channel had a significant number of artifacts, we then removed the entire channel from the training set of the SVM. A channel was dynamically excluded if the channel had more than 10% of non-neural artifacts in the 20-minute dynamic training set (explained in section 2.5.1). By taking these measures we ensured that the training data for the SVM contained only neural data. All extracted features were normalized before its training and classification by fitting the features to an arctangent function using the minimum and maximum values from the training set.

### 2.5 Support vector machines

Seizure detection involves the classification of ictal events in a highly unbalanced dataset, since ictal activity is highly uncommon in comparison to the amount of interictal activity. Then a one-class SVM was chosen to classify seizures as novel events by training the SVM on interictal data. The basic idea behind the one-class SVM is to map feature vectors to a higher dimensional space (i.e. feature space) using a kernel function, to maximally separate the data from the origin using a hyperplane. If a data point lies beyond the hyperplane it is considered a novel event. In this section we first describe the kernel function to then explain the SVM functionality and its implementation. The Gaussian kernel function was used throughout this study (Müller et al. 2001). This function is widely adopted in kernel methods (Xiao et al. 2015), since it can handle nonlinearities between feature vectors and its classification. The Gaussian kernel function is of the following form:

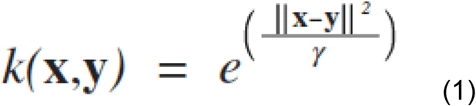

In Equation (1), the feature vectors **x** are mapped into a higher dimensional space called the feature space. The Gaussian width of the kernel function is given by *γ*, which is the parameter to be selected. The Gaussian width will determine the morphology of the closed curve surfaces that enclose the feature vectors to be mapped in the feature space. A suitable kernel parameter is supposed to accurately map the geometric distribution of feature vectors in the feature space to maximize classification performance (Xiao et al. 2015). Once feature vectors are mapped onto the feature space the one-class SVM constructs a hyperplane to linearly separate the data points at a maximal distance from the origin. The distance from any data point to this hyperplane entails the novelty of the datapoint with regards to the training data. The hyperplane parameters are then determined by solving the following quadratic programming problem (Gardner et al. 2006):

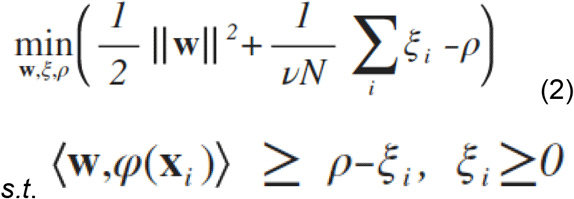

With the kernel function; *k(***x**_*i*_,**x**_*j*_) = (*φ (***x**_*i*_), *φ* (**x**_*j*_)). For the once-class SVM the only control parameter is *ν*, which is a tradeoff parameter between the model complexity and the training error. Thus, *ν* is an upper-bound on the fraction of data points lying on the wrong side of the hyperplane, and a lower-bound on the fraction of support vectors returned by the algorithm. Therefore, *ν* can be interpreted as the fraction of data points labeled as outliers, with this in mind we can fix its value to the seizure frequency observed in EEG data to analyze (Gardner et al. 2006). After estimating the fraction of ictal data in our recordings; a value of 0.01 was chosen for *ν*. This was calculated using the third quartile total duration of seizures (∼180 sec) as described by Afra et al. 2008. Consistent values for *ν* were obtained when calculating over the length of our entire dataset, and by patient.

#### 2.5.1 Dynamic training for SVM

Our innovative training paradigm of the one-class SVM, consists of using a new training set every second. The dynamic training set comprises the previous 20-minute window of features with a five-minute delay from the current instance. This was done to effectively separate and classify between interictal data and ictal data. With this delay we are able to successfully exclude preictal activity preceding the seizure from the training set. Figure 1 illustrates the dynamic training window for the SVM. This dynamic training approach yields an adaptive robust method that is unaffected by able to adapt to ultradian and circadian rhythms as well as to electrographical changes induced during sleep.

**Figure 1.**
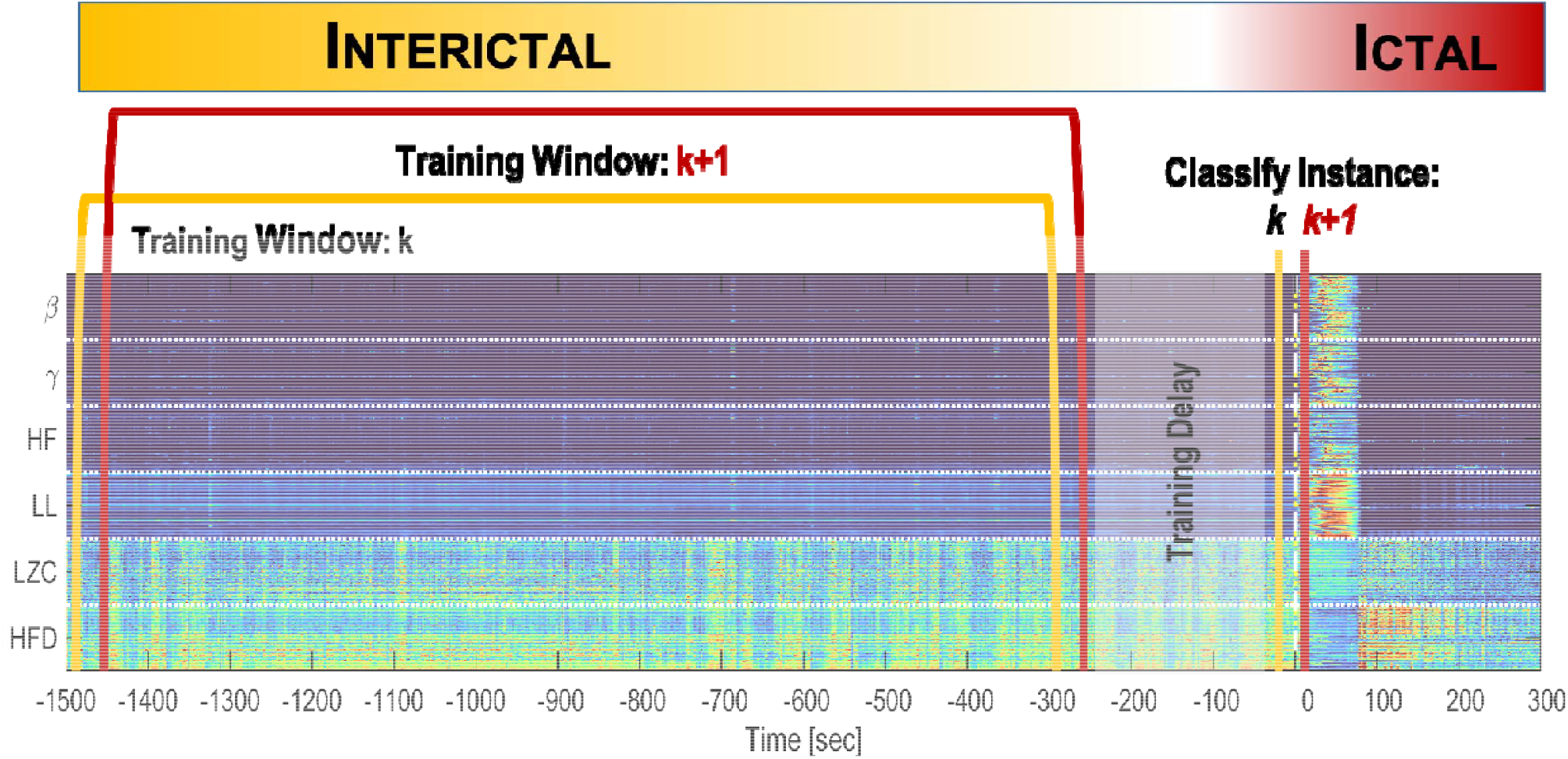
Dynamic training paradigm. Heatmap shows the extracted features from the EEG over time. Every second, a shifting 20-minute window of features is used for SVM training. This training window is delayed five minutes from the current instance to exclude any preictal data that might mislead the SVM in classification of the current instance.

#### 2.5.2 Multi-SVM architecture

In this study, we first implemented a one-class SVM for all channels and features as it has been used previously (Nandan et al. 2010; Park et al. 2011). We then compare the performance of this configuration with two multi-SVM configurations. Where a one-class SVM is assigned to determine novelty for (A) each channel location (i.e. strip, grid or depth electrode), and for each feature separately (B). For (A) and (B) the output is given by the number of SVMs that consider the current instance as novel.

### 2.6 Postprocessing

Each second the current feature vector was mapped onto the feature space and depending on its location with regards to the hyperplane its novelty was evaluated. The output of the SVM was then the distance to the hyperplane, the more negative the more novel that feature vector is considered. Since the computed distance to the hyperplane can often have noise, the use of filtering methods to increase detection specificity is necessary. A Kalman Filter was implemented on the output of the SVM, by using a *white noise acceleration* model to track the output of the SVM (Chisci et al. 2010; Park et al. 2011). This model assumes that the signal to track varies at a constant velocity, allowing the Kalman Filter to make the best prediction that minimizes the variance (i.e. noise, rapid changes) in the measured signal. The postprocessed output of the SVM is the signal used by our algorithm that triggers events for event detection of seizures.

### 2.7 Seizure detection

An ictal event was detected when the output of the algorithm crosses a threshold value. The post-processed output of the SVM denotes the smoothed distance to the hyperplane. In order to trigger a novelty event detection we used a predetermined threshold value on this output variable. The calculation of this threshold is described below in section 2.9. For multi-SVM configurations (A) and (B) an event is detected when a minimum of two SVMs or 20% of the total number of SVMs cross the predetermined threshold.

Once an event was detected, a refractory period was activated. The main purpose of this refractory period is to ignore any subsequent detections on the same event provoked by long seizures or postictal activity. During the refractory period the training of the SVM was paused, and is only resumed after the termination of the refractory period excluding this period from the training set. Afra et al. 2008 reported that seizures have a maximal duration of approximately 300 seconds with a third quartile value around 180 seconds. With this in mind we set the refractory period to be 300 seconds long. A nullification rule to the refractory period was added; if the output of the algorithm crosses the hyperplane to the positive side (nullifying all novelty in the current instance), then the refractory period is terminated and there is place for a new detection. This was implemented because as the algorithm is implemented in real-time, some seizures are not detected due to non-ictal detections within less than 300 seconds of seizure onset.

### 2.8 Performance evaluation

To evaluate the performance of the algorithm we used seizure onset markings determined by certified epileptologists (M.C.C. and G.K.B.). Detection windows of different sizes around the marked onset were used, to evaluate and track the performance of the algorithm for early-onset seizure detection and as a universal seizure detector. Sensitivity and specificity were calculated per patient and then averaged across patients.

#### 2.8.1 Early seizure-onset detection

It has been suggested that the window of opportunity to detect a seizure before clinical symptoms arise can be up to 30 seconds after electrographic onset (Hao Qu and Gotman 1997; Jouny et al. 2011). With this in mind, we defined an early true positive (TP) as an event detected 30 seconds before or 30 seconds after the clinically marked seizure onset. Any event detected outside this 60-second window is counted as a false positive (FP) for early seizure-onset detection. Sensitivity of the algorithm was estimated as the number of TP over the total number of seizures (TPr). The number of FP per hour (FP/hr) was evaluated as the algorithm’s specificity.

#### 2.8.2 Universal seizure detector

In order to assess the overall performance of the algorithm as a general seizure detector, we relaxed the constraint of using a specific window size of 60 seconds around the marked onset for early seizure-onset detection. We analyzed the performance of the algorithm with different sizes of detection windows besides the 60 second window mentioned in the previous subsection. The sizes of the windows are the following: [−45 +45], [−60 +60], [−60 +120] and [−60 +300], in units of seconds in relation to the marked onset. The relaxation of this parameter shows that even though the algorithm might miss an early-seizure onset detection, it still exhibits high performance in overall detection of seizure events.

#### 2.8.3 Non-Evolving Epileptiform Activity (NEEA)

Interictal recordings in patients with focal epilepsy occasionally contain short bursts (3-10 seconds) of epileptiform activity, or interictal discharges with spike-and-wave patterns (Gotman 2011). Given that our algorithm is intended for early seizure onset detection, it is sensitive to any sudden ictal or interictal epileptiform activity of significance. The emphasis on doing fast detection prevents the algorithm from confirming the evolution of the abnormal signature of interictal epileptiform activity into a seizure. We conducted a post-hoc review of the recordings of six patients to identify those events and assess their impact on overall performance of the algorithm to determine the impact of NEEA events on the algorithm efficiency.

### 2.9 Parameter optimization

Given that our algorithm uses a dynamic training set of features that is changing every second, a grid search of the event-detection threshold value and the Gaussian width value (*γ*) was performed to optimize the parameters. The grid search was performed on the same random set of seizures used for calibration of the artifact detector. Each seizure was contained in a two-hour EEG epoch. Threshold values ranged from 0 to −5 in steps of −0.5, Gaussian width values ranged from 0.001 to 0.01 in steps of 0.001. The range of values explored for *γ* was determined by calculating the mean Euclidian distance within the training set as well as using the local geometric information of the feature vectors as described by Xiao et al. 2015.

The largest threshold with 90% sensitivity was picked. This was done, as it is logical to assume that as the threshold increases; the amount of FP decreases, detection latency increases and sensitivity is lost. Then, by picking the highest threshold with a 90% sensitivity constraint, we maximize the specificity for that performance.

### 2.10 Statistical analysis

We used a two-factorial nonparametric test (Scheirer-Ray Hare) to assess significance of the performance metrics (i.e. TPr, FP/hr and latency) among the different SVM configurations. The Scheirer-Ray-Hare test was used to compare results metrics of the three SVM configurations (single-SVM and multi-SVM configurations; i.e. A and B), and the five different detection window sizes ([−30 +30], [−45 +45], [−60 +60], [−60 +120] and [−60+300]). Following the rejection of the null hypothesis, the Dunn’s test was used as a post-hoc test to compare performance differences among multiple groups. The default alpha value was 0.05.

## 3. Results

A dynamic learning SVM algorithm for early-onset seizure detection was developed with the goal of being implemented for real-time detection of seizures in the EMU. Detection performance was evaluated for real-time early seizure-onset detection in the EMU using one and multi-SVM configurations. Performance of the algorithm as a universal seizure detector was assessed using different detection window sizes. Finally, a partial review of six patients was done to analyze the impact on post-hoc assessment of FP events.

### 3.1 Detection performance for early seizure-onset detection

Detection of early seizure-onset used a detection window of 30 seconds before and after the marked onset of the seizure by clinicians. An example of a successful early-onset seizure onset detection is shown in Figure 1, and a late detection is shown in Figure 2. The best results were achieved using a single-SVM configuration, with a TPr of 87%, a FP/hr of 1.25, and a mean detection latency of 10.4 seconds. When using multi-SVM configurations A and B as described above in section 2.5.2, TPr was 77.9%, and 78.7% respectively. FP/hr was 0.86 for configuration A and 1.72 for B. Detection latencies were 13.3 and 11.7 seconds, for configurations A and B. These results are shown in the first column of Table 1 and Figure 4.

**Figure 2.**
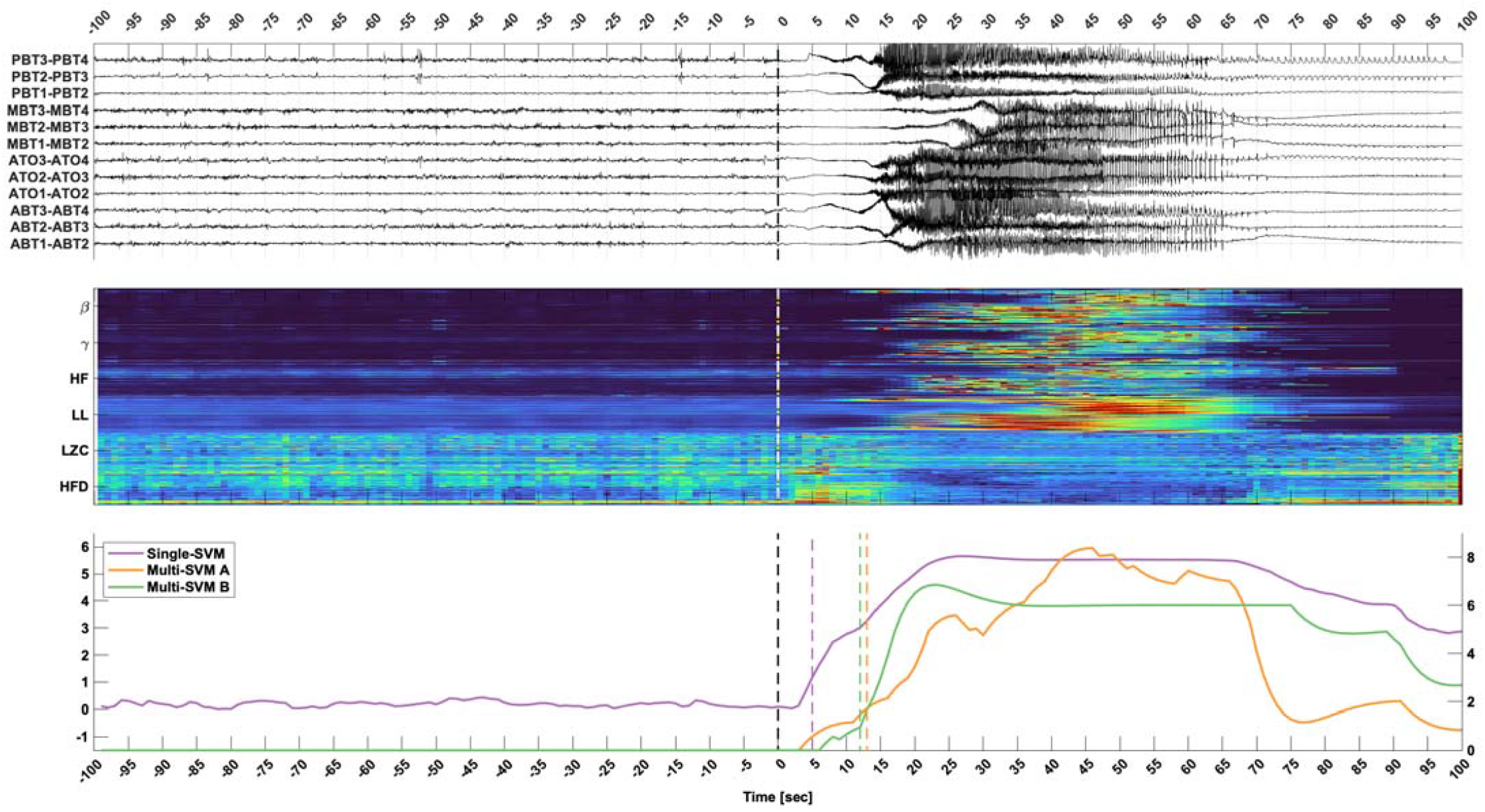
Early-onset detection. Top panel shows 200 seconds of the EEG of a selected number of channels where the seizure onset zone is localized. Middle panel shows the extracted features from the EEG. Bottom panel shows the postprocessed output for the different SVM configurations. The purple trace represents the single-SVM, the orange trace the multi-SVM A, and in green the multi-SVM B configuration, respectively. Dashed colored lines show when the event was detected for each SVM configuration. The dashed black line at time zero denotes the seizure onset as marked by the clinicians.

**Figure 3.**
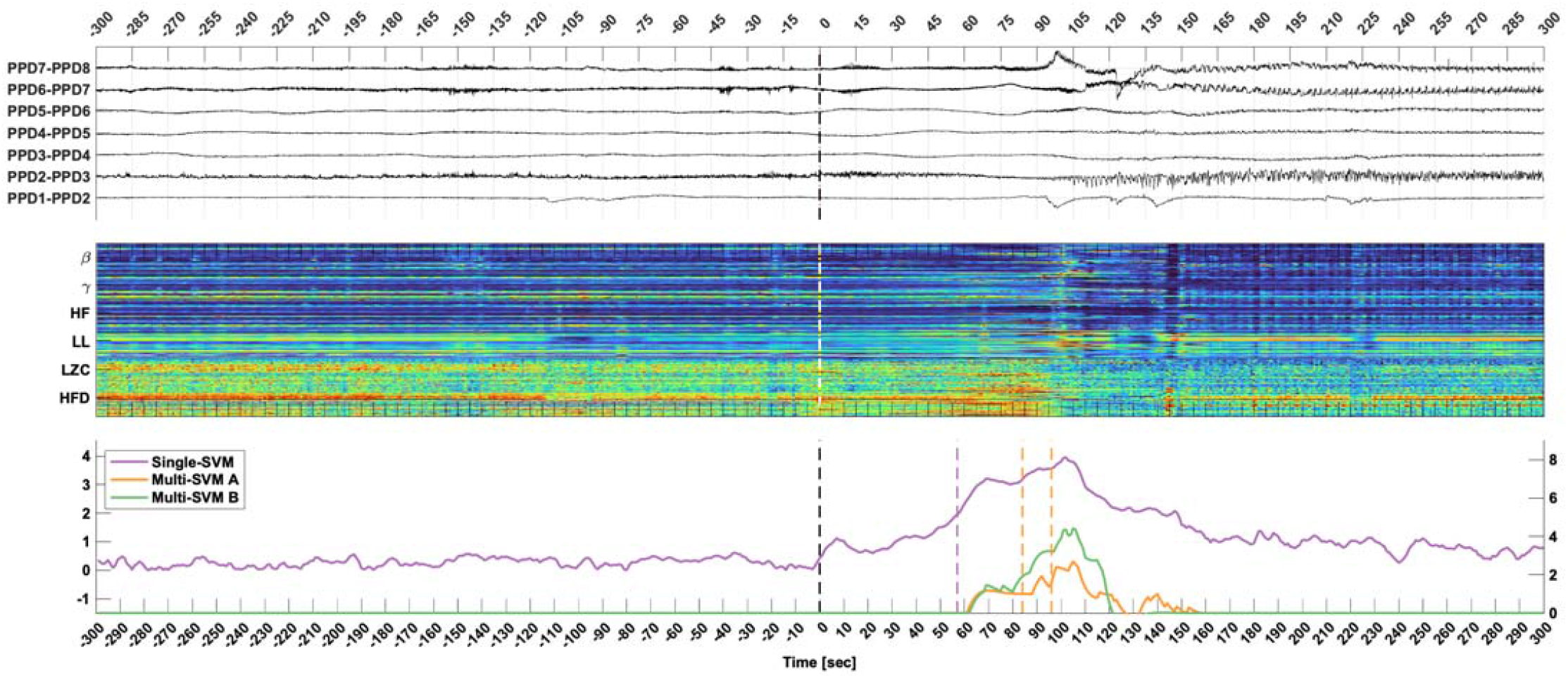
Late detection. Top panel shows ten minutes of EEG signal for a depth electrode that shows maximal signal change during seizure. Middle trace shows the extracted features from the EEG. Bottom trace shows the postprocessed output of the SVM, color-coded as in Figure 2. Vertical dashed colored lines show when the event was detected for each SVM configuration respectively. The dashed black line at time zero denotes the seizure onset as marked by the clinicians.

**Figure 4.**
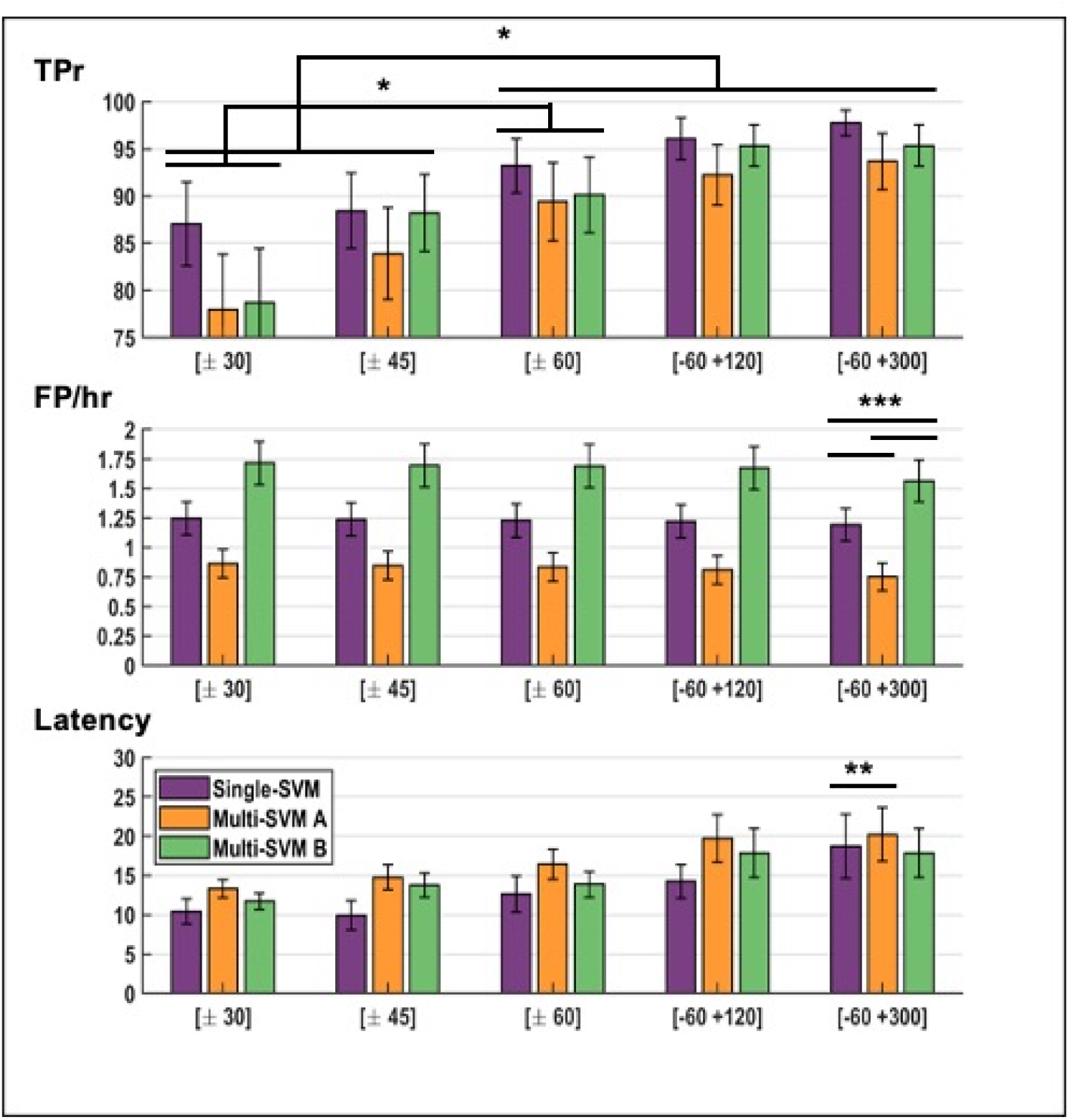
Results Stats in bar plots. The results are shown for TPr, FP/hr and Latency for the different SVM configurations as well as for the different detection window sizes. Statistical significance is shown for post-hoc analysis using Dunn’s test (* p <= 0.05, ** p <= 0.001, and *** p < 0.0001).

**Table 1.**
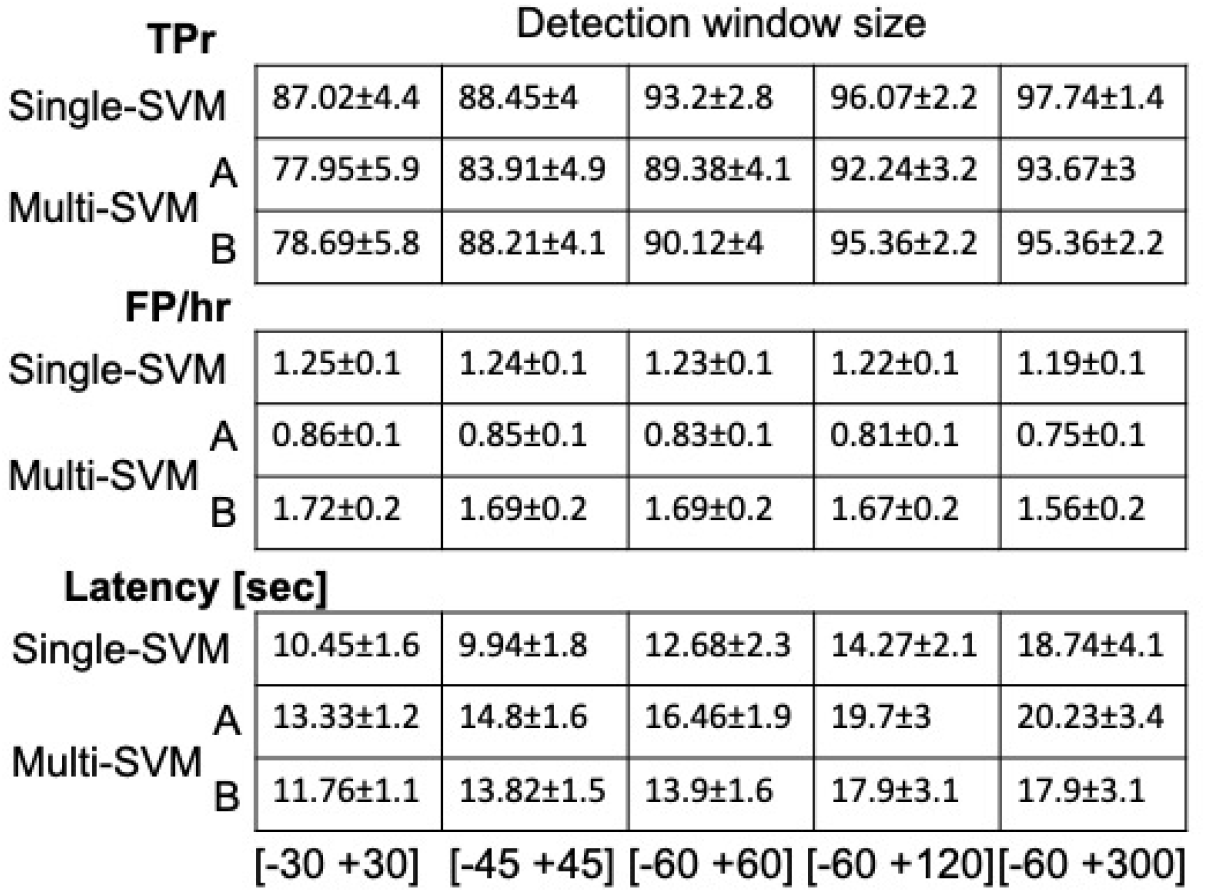
Detection results showing the TPr, FP/hr and latency for the different SVM configurations as well as for the different detection window sizes.

### 3.2 Performance for different detection window sizes

In order to evaluate the performance of our algorithm as a generic seizure detector and help clinicians detect and localize seizures, we used different detection windows sizes around the clinically marked seizure onset, as described in section 2.8. Results for the different windows are presented in Table 1. The increase in detection window size had a positive effect on performance metrics for all SVM configurations; increasing the TPr, decreasing the FP/hr. However, detection latency was increased as an expected trade-off. For the largest detection window size (i.e. [−60 +300]) single-SVM and multi-SVM configurations (A and B), reached an overall sensitivity of 97.7%, 93.7% and 95.4% respectively. FP/hr was decreased to 1.19, 0.75, 1.56 accordingly. Finally, latency was increased to 18.7 seconds for the single-SVM configuration. For multi-SVM configurations A and B, mean detection latency values were 20.2 and 17.9 seconds respectively. Performance results for different SVM configurations and detection window sizes are shown in Figure 4.

### 3.3 SVM configuration and detection window size effects on detection performance metrics

Nonparametric two-factorial statistical analysis on detection results using the Scheirer-Ray-Hare test showed the effect of varying the detection window size and using different SVM configurations. For TPr, the detection window size showed an influence on detection results, rejecting the null hypothesis (p = 0.0005). Post-hoc analysis using the Dunn test for TPr shows significance between several window sizes as illustrated in Figure 4. Regarding FP/hr, the SVM configuration had a strong statistical significant effect among all configurations (single-SVM, multi-SVM A and B) for all window sizes, with post-hoc p values under five decimal values. Finally, for detection latencies the SVM configuration also played a significant role (p = 0.001) for all window sizes. Post-hoc analysis showed a significant difference between single-SVM and multi-SVM configurations, p = 0.001.

### 3.4 NEEA assessment

Our algorithm was also sensitive to NEEA, which are helpful for surgical evaluation and seizure characterization. An exemplary detection of a NEEA is illustrated in Figure 5. To assess the impact of FP events detected by our algorithm in our recordings, and whether these events were triggered by non-neural artifacts or NEEAs a partial review of results was done on 6 patients. For these patients we used two detection window sizes (i.e. [−60 +60] and [−60 +300] seconds) for seizure detection. This was done to evaluate the impact on TPr and FP/hr performance metrics. Subsequently, a [−60 +60] second window size was used to evaluate triggered FP. After careful revision of FP detected events and labeling of NEEAs we recomputed the TPr and FP/r. Results showed that for the [−60 +60] detection window, TPr = 89.6% before addition of NEEAs, and then increased to 96.3% with NEEAs. Conversely, FP/hr decreased from 0.83 to 0.41. For the largest detection window, [−60 +300], TPr increased from 97.7% to 99.6%, and FP/hr decreased from 0.81 to 0.38.

**Figure 5.**
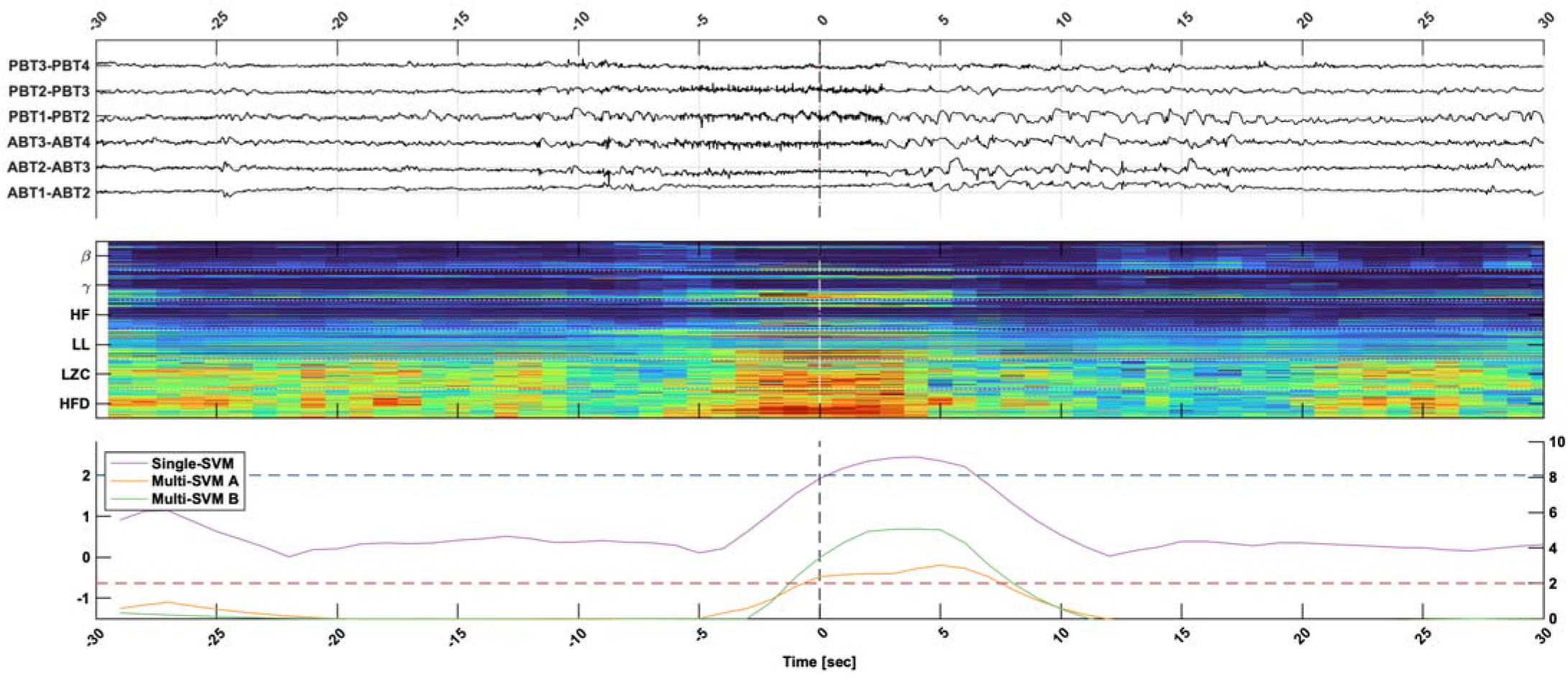
NEEA detection. Top panel, shows one minute of EEG trace, for subset of channels that show epileptiform activity. Middle panel, shows extracted features from the EEG. Bottom panel as in Figure 2 and 3 shows the postprocessed output of the different SVM configurations. Horizontal dashed lines show the detection threshold for the SVM configurations. In blue the threshold for the single-SVM and in red the threshold for the multi-SVM configurations. The dashed black line at time zero denotes the NEEA event marked by the clinicians.

## 4. Discussion

There is a clear immediate need for a warning system designed for early seizure-onset detection in the Epilepsy Monitoring Unit (EMU) that is reliable and efficient for clinical staff to provide immediate care to patients with epilepsy during seizures and ensure their safety (Carlson 2009; Noe and Drazkowski 2009; Jouny et al. 2011; Atkinson et al. 2012; Lee and Shah 2013).

Therefore, such a warning system must rely on an algorithm that is computationally efficient and can be easily implemented in real-time. Additionally, it must be sensitive to a wide range of seizure-onset patterns and still maintain high specificity to avoid alarm fatigue (Fürbass et al. 2015; Kamitaki et al. 2019; Bi et al. 2020).

Modern algorithms and current technological systems allow for the development of practical and efficient warning system, however, this need remains unmet. Many of the recently developed algorithms are not practical for implementation as an alarm system that is sensitive to early detection of seizures in the EMU (Stacey 2018; Kamitaki et al. 2019). Several algorithms have been developed for early-seizure-onset detection with real-time capabilities for scalp-EEG (Meier et al. 2008a; Chisci et al. 2010; Minasyan et al. 2010; Fürbass et al. 2015; Bogaarts et al. 2016; Sridevi et al. 2019). However, these algorithms have not been validated for intracranial EEG recordings. Patients admitted to the EMU that undergo intracranial EEG recording are at a higher risk of serious injury (e.g. unintended electrode extraction or intracranial hemorrhage due to head injury during seizure). Furthermore, the number of electrodes utilized increases significantly (on average, four-fold in our dataset) with the use of intracranial recordings, compared to scalp EEG. The increase in number of recording contacts reduces the signal-to-noise ratio which can affect the performance of detection algorithms.

More modern algorithms that analyze intracranial EEG recordings have implemented machine learning techniques as these allow for the selective learning of features relevant to ictal activity to train classification models. Machine learning algorithms have recently shown great potential for practical seizure prediction (Acharya et al. 2018; Kiral-Kornek et al. 2018; Kuhlmann et al. 2018; Tsiouris et al. 2018; Abbasi and Goldenholz 2019). These algorithms are able to classify electroencephalographic features into interictal, ictal and in some cases preictal data. However, the performance of these algorithms depends on their training dataset. The use of these multi-class algorithms that are trained on crowd-sourced data, presents a risk of overfitting and/or requires an exceptionally large training dataset to capture the entire feature space of ictal activity. This is particularly difficult due to the complexity of seizure dynamics, the large variability of seizure-onset patterns and the localization of the seizure onset focus in relation to the implanted electrodes, which varies between patients.

Personalized medicine and patient-specific algorithms provide many advantages, as they increase the sensitivity and specificity of detecting the patient’s signature ictal patterns taking into account electrode coverage (Hao Qu and Gotman 1997; Kharbouch et al. 2011; Santaniello et al. 2011; Tsiouris et al. 2018). However, this is only feasible for chronic implantation of electrodes (i.e. RNS), where a database of ictal activity is being built continuously for each patient and used to train the algorithm. This is not viable for a seizure-alarm system in the EMU, where prerecorded seizures are not available at the time of patient presurgical evaluation.

In this study, we propose a patient-specific and self-adaptive intelligent algorithm that is able to analyze long EEG recordings and quickly classify ictal activity as novel to interictal activity. Our algorithm is based on the use of a one-class SVM (Gardner et al. 2006). This powerful algorithm is able to classify the level of novelty with respect to its training data, detecting abnormal events. The principle of this algorithm is to detect seizure as abnormal events when trained with interictal data(Figure 6). The computational efficiency of the algorithm allows for easy implementation in the EMU as real-time alarm system for detection of early seizure-onset patterns.

**Figure 6.**
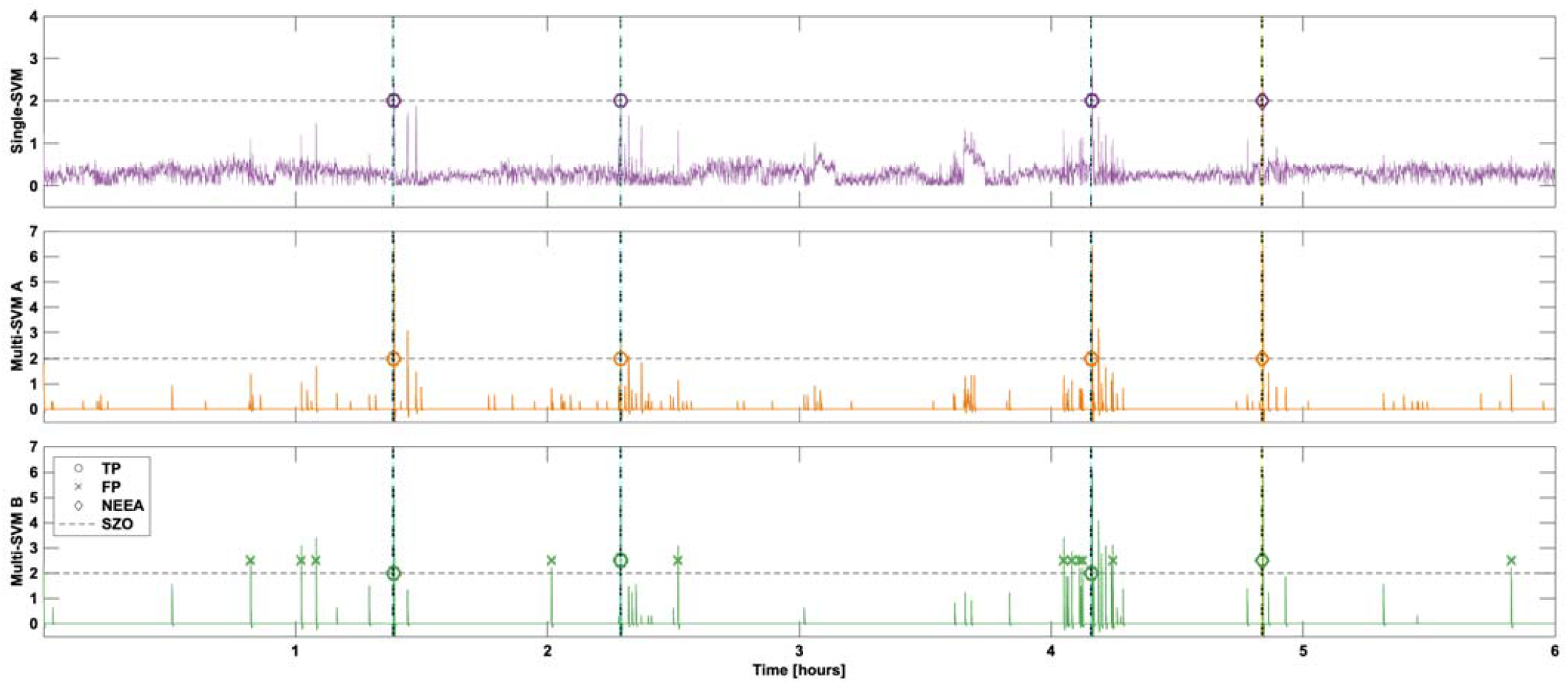
Long SVM output traces showing six-hour traces of the SVM output for all three configurations, single-SVM (purple), Multi-SVM A (orange), and Multi-SVM B (green). Seizure-onset markings are show in vertical dashed black lines. Detection thresholds in horizontal dashed lines. TP detections are marked with a circle, FP are marked with a cross, and NEEA detections are marked with a diamond.

With one-class SVM used for novelty classification, we introduce an innovative dynamic training framework that does not rely on prior ictal data. This algorithm therefore has several benefits: i) no a priori data is needed for it to become a patient-specific algorithm. ii) due to its adaptive nature it is not affected by the slow variation in EEG patterns linked to ultradian and circadian rhythms. Our dynamic training paradigm consists of a continuously shifting training window of past interictal features sensitive to ictal activity as shown in Figure 1. This training window has a five minute delay from the current instance to exclude any preictal activity before seizure onset that might mislead the novelty classifier (Litt et al. 2004).

We considered any detection within a 30 second window of the clinician marked seizure onset to be an early seizure-onset detection. This window of opportunity after the electrographic seizure onset and before clinical symptoms arise is crucial for a warning system to alert clinical staff (Hao Qu and Gotman 1997; Jouny et al. 2011). Our algorithm achieved a high-performance for early seizure-onset detection, reaching a TPr of 87% with a FP/hr rate of 1.25 and a mean detection latency of 10.45 seconds. An example of an early seizure-onset detection is shown in Figure 2.

Our detection results are summarized in Table 1 and in Figure 4. We compared three different SVM configurations; using a single-SVM for all channels and features, multiple SVMs by channel location (multi-SVM A and for each feature multi-SVM B). This was done to increase the specificity and/or sensitivity of the algorithm in case a patient had only some channel locations recording early ictal patterns. Alternatively, specific features (e.g. HFD and power in the β band) could be more sensitive to detecting a particular patient’s ictal patterns, as shown in previous studies (Jouny and Bergey 2012). Using multiple one-class SVMs and isolating sensitive features for training of each SVM, increases the sensitivity and/or specificity to a specific subset of the feature space. For the multi-SVM A, it increased the specificity by decreasing the FP/hr rate. And for the multi-SVM B it showed an increased sensitivity in comparison to configuration A, but not in comparison to the single-SVM. Results in Table 1 also show the superiority of the single-SVM compared to the multi-SVM configurations for early seizure-onset detection. Other multi SVM configurations were initially tested with no significant results over the ones described here. We tried the usage of an SVM per feature and per channel location together (combining configuration A and B) as well as the usage of a second layer to determine novelty over the output of the first layer of SVMs. There was no significant improvement relative to the increase in complexity of the processing to warrant pursuing those configurations.

Further comparison between the different SVM configurations from Table 1, shows that the single-SVM configuration had overall better sensitivity although not statistically significant against the other configurations. The multi-SVM A configuration showed the lowest FP/hr ratio and the multi-class SVM B had the largest FP/hr ratio, twice as large as the multi-SVM A. For detection latency, the single-SVM had the best performance and statistical significance over the multi-SVM A (See Figure 4). We concluded that the highest performance configuration is achieved by the single-SVM configuration.

It is worth noting from that the average detection latency for the single-SVM is 10.4 seconds for our real-time seizure onset detector. Apart from patients with onset of seizures originating from primary motor areas, we anticipated this detection to occur before clinical symptoms arise. It is important to notice from our results in Table 1 that the mean latency and standard deviation even for the largest detection window size fell within our early seizure-onset window of opportunity.

The single-SVM for early seizure-onset had a 100% sensitivity in 74% (26/35) of the patients studied. At least one seizure was detected for 97% (34/35) of the patients. As shown in Figure 3, some seizures have a very slow evolving seizure onset. This would potentially mean that even a triggered detection 30 seconds following electrographic seizure onset might be useful and precede the onset of clinical symptoms.

We also evaluated our algorithm as a universal seizure detector. When using a detection window of [−60 +60] or larger, the algorithm had a true detection for at least one seizure in all patients.

Our results show a high sensitivity for detection of epileptiform activity, be it a late detection or Non-Evolving Epileptiform Activity (NEEA). When using the largest window detection interval ([−60 +300]) the algorithm achieves a sensitivity of 97.7%. It is important to mention, that in this case it could be that the seizure is terminated before the 300 second detection window, and that the detection was triggered by postictal activity which would also be classified as novel compared to baseline activity.

However, if the seizure was detected well after the onset, this could warn the staff to assess the patient during a prolonged seizure, before a seizure progresses to a bilateral tonic-clonic seizure, or post-ictally Moreover, when the clinical staff becomes aware that a patient is having a seizure they interact with the patient during a seizure to test for cognitive skills to better characterize the evolution of the seizure. An efficient alarm system offers many advantages as a technological tool for clinical staff in the EMU to offer better care to patients regardless if the alarm not always precedes the onset of clinical symptoms. The high overall-sensitivity to seizures shows that this algorithm can help expedite the EEG-review process that clinicians go through to find seizures and characterize them which has been traditionally done by visually analyzing hundreds of hours of EEG in a fast-forward manner. In addition, our false positive rate is well within an acceptable range as a warning system to avoid alarm fatigue (Lee and Shah 2013; Tovar Quiroga et al. 2016; Bi et al. 2020). The false positive rate still remains higher in comparison to other algorithms (e.g. Fürbass et al. 2015), however, our algorithm is designed for fast detection, therefore it cannot distinguish the evolution of epileptiform activity.

To explore this we performed a post-hoc analysis of the triggered events for six patients, we realized that some events labeled as FPs were in fact NEEA, which is nonetheless helpful for characterization of the seizure onset zone for presurgical evaluation. Often this NEEA is not a false-positive detection as true epileptiform activity is detected, but represent events that do not evolve into clinical seizures (Jouny et al. 2011; Beniczky and Ryvlin 2018). Despite their short duration, the dynamics of these events often possess characteristics identical to those of disabling clinical seizures. Undoubtedly, these events are helpful for surgical resection planning. An example of a detected NEEA is shown in Figure 5. There were a total of 54 NEEAs marked for the 6 patients with a mean 9 NEEAs per patient. For these patients the early-seizure onset detection sensitivity increased significantly from 89.6% to 96.3% paired with a decrease in FPr/hr from 0.83 to 0.41.

The definition of what a seizure is, and what constitutes a seizure remains a topic of debate (Fisher et al. 2005). The lack of a clear definition for seizures inevitably hinders objective meta-analysis of seizure detection algorithms and obscures their true performance. Here we presented an algorithm that is patient-specific, robust, practical to implement in the EMU and sensitive to epileptiform activity. This algorithm is a potentially valuable tool for clinical staff in the EMU; as an early seizure detection alarm system, and a universal detector of ictal activity as well as any epileptiform activity that could be useful for surgical planning.

## 5. Conclusion

This study presents a robust self-adaptive algorithm that is able to detect seizures reliably without prior training and is practical to implement in the EMU. Our algorithm renders a computational tool that achieved high performance for early-seizure onset detection which could be utilized clinically as a warning system in the EMU. It also proved helpful for seizure characterization and can expedite the review process of ictal events in long-term EEG recordings for clinicians.

## Data Availability

Data is not available.

## Acknowledgements

Research supported by NIH R01 – NS75020. Daniel Ehrens is a Howard Hughes Medical Institute Gilliam Fellow.

